# The association between academic achievement goals and adolescent depressive symptoms: a prospective cohort study in Australia

**DOI:** 10.1101/2023.11.02.23297905

**Authors:** Thomas Steare, Glyn Lewis, Katherine Lange, Gemma Lewis

## Abstract

**Background:** Students define competence according to the development of their skills and understanding (mastery) or based on comparisons with their peers (performance). Students may aim to achieve the outcome of interest (approach) or to avoid failure of not meeting their definition of competence (avoidance). Achievement goals are linked to adolescents’ cognitions, coping, stress, and potentially depressive symptoms. We conducted the first longitudinal study of the association between achievement goals and depressive symptoms in a nationally representative adolescent sample.

**Methods:** We analysed data from the Longitudinal Study of Australian Children. Achievement goals were measured at age 12/13, with the primary outcome (depressive symptoms) measured at ages 14/15 and 16/17 (Kindergarten only). Analyses were linear multilevel and traditional regressions, with adjustment for confounders.

**Outcomes:** We included 3,200 and 2,671 participants from the Kindergarten and Baby cohorts, respectively. Higher mastery-approach goals were associated with decreased depressive symptoms (Kindergarten: −0·33 [95%CI: −0·52 to −0·15]; Baby: −0·29, [95%CI: −0·54 to −0·03]), and higher mastery-avoidance with increased depressive symptoms (Kindergarten: 0·35, [95%CI: 0·21 to 0·48]; Baby: 0·44 [95%CI: 0·25 to 0·66]). Higher performance-avoidance goals were associated with increased depressive symptoms in the Kindergarten cohort only (Kindergarten: 0·26, [95%CI: 0·11 to 0·41]; Baby: −0·04 [95% CI: −0·27 to 0·19]). We found little evidence of an association between performance-approach goals and depressive symptoms.

**Interpretation:** If associations reflect a causal relationship, school environments that promote mastery-approach goals, could reduce adolescent depressive symptoms.

**Funding:** Sir Henry Dale Fellowship jointly funded by the Wellcome Trust and the Royal Society (Grant 223248/Z/21/Z).

**Research in context:** *Evidence before this study:* The onset of depression commonly occurs in adolescence, with evidence suggesting that the rate of adolescent depression is rising in many countries. Despite the importance of prevention, very few strategies are successful. Modifications to the school environment may lead to improvements in adolescents’ mental health, however there is little awareness over which factors to target. Achievement goals reflect differences in adolescents’ motivation to learn and how they measure their own success, have been linked to adolescents’ cognitions, ways of coping, stress, anxiety, and self-esteem. According to the goal-orientation model of depression vulnerability, they represent a potential risk factor for adolescent depression. Evidence has shown that students achievement goal orientations are influenced by the school environment, and that they are modifiable through targeted intervention. If there is a causal relationship, changing adolescents’ achievement goals may reduce depressive symptoms, however there is a lack of high quality evidence. We searched MEDLINE and PsycInfo from database inception to August 1, 2023 for studies describing the association between achievement goals and depression in adolescents. We used the search terms “achievement goal*” AND “adolescen*” OR “student*” AND “depression”, alongside searches in Google Scholar. We found only one longitudinal study, which used an unrepresentative convenience sample and did not adjust for confounders.

*Added value of this study:* Using longitudinal data of two nationally representative Australian cohorts, we examined the association between four achievement goal orientations with subsequent depressive symptoms in school-going adolescents. We found longitudinal evidence that mastery-approach, mastery-avoidance, and performance-avoidance goals were moderately associated with depressive symptoms independent of a range of confounders.

*Implications of all available evidence:* Our study highlights that adolescents’ cognitions around learning and success are associated with future mental health outcomes. School environments that promote learning, development and personal growth could therefore reduce adolescent depressive symptoms, if this association were causal. School-based interventions that aim to enhance factors consistent with mastery goals (i.e., learning skills and understanding the subject, rather than assessing competence in comparison to peers) may have success in preventing depression, and trials are therefore warranted.

## Introduction

Depression is among the top five contributors to the global burden of disease and predicted to be the leading cause of disability in high-income countries by 2030.^1^ Incidence increases sharply during adolescence ^2^, and evidence suggests that rates of adolescent depression are rising.^3^

Schools are a potential setting for preventative interventions that would reach most adolescents, and increase equity of access. Most school-based interventions to prevent or reduce mental health problems have used psychological approaches aimed at students as individuals (for example mindfulness, cognitive behavioural therapy, or psychoeducation).^4^ There is evidence that these approaches have generally been ineffective at reducing depressive symptoms.^4^ An alternative approach is to modify the school environment. For example, whole-school interventions aim to change the school culture, climate, and values (goals, norms, teaching practices and organisational structures).^5,6^ There is evidence from randomised trials that adolescent depressive symptoms can be reduced through whole-school interventions targeting health promotion and socio-emotional skills.^7,8^ Many whole-school interventions are unsuccessful however, and we need a better understanding of which risk factors to target.^9^

Achievement goals have been widely studied in research on motivation, particularly within schools, and may represent a modifiable risk factor for adolescent depressive symptoms. Achievement goal orientations can be defined as cognitive representations that guide behaviour to an end state. In educational settings, they reflect differences in individuals’ motivation to achieve outcomes.

There are two key dimensions that differentiate achievement goals.^10^ The first is how someone defines competence and success for themselves. Mastery goals are when students define their success as developing their understanding or skills. In contrast, performance goals are when success is defined by out-performing peers. The second dimension refers to valence, meaning whether someone’s achievement goals are focused on either attaining success or avoiding failure. Mastery-approach goals therefore refer to the motivation to develop competence, task, or subject understanding, and to learn as much as possible. Mastery-avoidance goals refer to the motivation to avoid incompetence, and are characterised by concerns about not being able to learn or understand subject material.^11^ Performance-approach goals refer to the motivation to out-perform peers whereas performance-avoidance refers to the motivation to avoid appearing incompetent and performing worse than peers.

The goal-orientation model of depression vulnerability proposes that achievement orientations may be linked with vulnerability to depression.^12,13^ The model proposes that high levels of performance orientation goals, lead to external standards for success based on social comparisons, which may lead to increased stress, anxiety, and low self-esteem. The model also proposes that mastery-approach reduces vulnerability to depression. Mastery-orientation encourages adolescents to view challenges and setbacks as opportunities for learning and growth, leading to more adaptive ways of coping with stress or failure.

There is evidence that achievement goal orientations are modifiable using interventions aimed at the school environment, including whole-school approaches.^14–16^ Students’ achievement goal orientations are thought to be influenced by whether schools emphasise learning and personal growth (i.e., mastery goals) or competition and social comparison (i.e., performance goals).^15^ Interventions that aim to alter achievement goal orientations have therefore been aimed at school policies, teaching practices and organisational structures, to create environments that promote mastery goals and de-emphasize performance goals ^15^. Randomised trials have found that achievement goal interventions are associated with improvements in factors potentially associated with depressive symptoms. These factors include anxiety, physical activity, enjoyment of tasks and confidence/competence.^17^ School environments that emphasise mastery goals might also be associated with lower levels of academic pressure and perfectionism, potential risk factors for depressive symptoms.

To our knowledge, only one study has investigated the longitudinal association between achievement goals and future depressive symptoms. Adolescents with lower levels of mastery goals and higher levels of performance goals at age 13/14 were more likely to experience consistently high levels of depressive symptoms throughout adolescence.^18^ The study however did not control for any confounders and used an unrepresentative convenience sample of students from local schools in Helsinki, Finland. Consequently, there is limited understanding as to whether achievement goal orientations are a potential risk factor of adolescent depression.

We investigated whether adolescent achievement goal orientations were associated with subsequent depressive symptoms, using a large nationally representative Australian cohort. We hypothesised that mastery-approach goals would be associated with lower levels of subsequent depressive symptoms whereas mastery and performance-avoidance goals and performance-approach goals would be associated with higher levels of future depressive symptoms.

## Methods

### Study design and participants

The Longitudinal Study of Australian Children (LSAC) include two cohorts; the Kindergarten cohort recruited at age 4-5 and the Baby cohort, recruited at age 0–1· Both studies began in 2004, to investigate factors influencing childhood development. A total of 4,983 children (Kindergarten cohort) and 5,107 babies (Baby cohort) were recruited to the first wave, using a random selection of households from a random selection of 330 Australian postcodes. Sampling was stratified to ensure proportionality to the population of children in these areas. Data collection was primarily conducted every two years resulting in eight waves as of 2020, with a further two waves conducted a year apart during the COVID-19 pandemic. Data were collected from the index child, parents, carers, and teachers. Further details are described elsewhere.^19^

Our analyses consisted of adolescents that were registered to attend school at the time of the exposure, with those home schooled or not registered excluded. We used data from waves prior to the COVID-19 pandemic given the impact the pandemic had on adolescent mental health which may have interfered with potential associations between achievement goals and depressive symptoms.

#### Outcome

Adolescents reported depressive symptoms using the Short Mood and Feelings Questionnaire (sMFQ). The 13-item sMFQ assesses the severity of depressive symptoms in the previous two weeks. Scores range from 0 to 26, higher scores indicating more severe depressive symptoms. In the Kindergarten cohort, we used depressive symptoms as a repeated measures outcome at ages 14/15 and 16/17· In the Baby cohort, we used depressive symptoms at age 14/15 as the outcome (the last time-point depressive symptoms were measured in this cohort pre-pandemic).

Psychological distress was measured in the Kindergarten cohort, at age 18/19, with the 10-item Kessler Psychological Distress Scale (K10)^20^. Scores range from 10 to 50, with higher scores indicating more severe distress. The K10 has shown excellent specificity and sensitivity for depression diagnosis, including in a sample of Australian adolescents.^21^

#### Exposure

Achievement goals were measured using the 12-item self-reported Achievement Goal Questionnaire (AGQ),^10^ based on the two-by-two formulation of achievement goal theory. The AGQ was completed at age 12/13 in each cohort, and again at age 16/17 in the Kindergarten cohort (Supplementary Table 1). Primary analyses used the AGQ at age 12/13 in the Kindergarten and Baby cohorts.

The AGQ has four 3-item sub-scales, which have been found to represent distinct internally consistent achievement goals: mastery-approach, mastery-avoidance, performance-approach and performance-avoidance. Each item was rated on a 7-point Likert scale (1 – “not at all true of me” and 7 – “very true of me”). Example items include: “I want to learn as much as possible this year” (mastery-approach), “I am definitely concerned that I may not learn all that I can this year” (mastery-avoidance), “It is important for me to do well compared to other students this year” (performance-approach) and “the fear of performing poorly is often what motivates me” (performance-avoidance) . Each sub-scale has a possible score from three to 21, with higher scores indicating higher levels of each achievement goal.

#### Potential confounders

Confounders were selected based on existing studies and theoretical assumptions. Confounders were measured during the same wave as the exposure and included sex, maternal depressive symptoms (measured by the K-6), whether the study child was living in a single-parent household, and a composite variable regarding socio-economic position (Supplementary Figure 1). For both cohorts, we adjusted for baseline depressive symptoms measured with the sMFQ.

Linked data was obtained from the National Assessment Program – Literacy and Numeracy (NAPLAN). The NAPLAN contains scores on students numeric and literacy ability from a national assessment for grades 3, 5, 7 and 9· As scores were not available in each domain for all participants, we created an average score across each of the different assessment (reading, writing and, spelling, grammar and punctuation, and numeracy) to indicate academic achievement. We used data for grade 7, when students were aged 12 to 13·

#### Statistical analysis

All analyses were conducted using Stata 17. Our sample consisted of those with complete data on exposure, confounders, and at least one depressive symptoms outcome score. Sample weights were used in all analyses, except in our secondary analyses using mixed-effects logistic regression with depression as a binary outcome variable due to issues with non-convergence.

##### Primary analysis

To model the repeated outcomes in the Kindergarten cohort, analyses were linear multilevel regression models with depressive symptoms at follow-up clustered within participants, and a random intercept for participant. In the Baby cohort, analyses were traditional linear regression models as there was only one follow-up time-point with the sMFQ. We first investigated univariable associations between each AGQ sub-scale separately and depressive symptoms. Next, we added all AGQ sub-scales (as correlations between them were lower than 0·7) (Supplementary Table 2). For multilevel regression models, we added a variable indicating follow-up time-point. We then added baseline depressive symptoms. Next, we added all other confounders. In the Kindergarten cohort analyses, we investigated whether associations between AGQ subscales and depressive symptoms differed according to follow-up, using an interaction between each AGQ sub-scale and time.

##### Secondary analysis

In the Kindergarten cohort, we also investigated associations with K10 scores as the outcome. This allowed us to test whether associations replicated with an older age group (18/19 years) and a different outcome measure. We investigated associations of AGQ scores at age 12/13 and 16/17, with the K10 at age 18/19 using linear regression models and a similar model building procedure to the primary analysis.

##### Sensitivity analyses

First, we included scores from NAPLAN assessments as an additional confounder in our primary analyses of each cohort. Grade 7 NAPLAN assessments took place after the exposure time-point for 9·1% and 12·4% of the Kindergarten and Baby cohort samples respectively, and therefore were not included in main analyses. Second, we conducted logistic multilevel modelling for the Kindergarten cohort with a binary depression outcome (SMFQ score of 8 or higher)^22^ using the same modelling procedure described above. For the Baby cohort, we ran logistic regressions with a binary depression outcome at age 14/15·

Third, we conducted multiple imputation to replace missing data. We imputed missing data in exposure, outcome, and confounders across the whole LSAC cohorts (Kindergarten cohort: n = 4,983; Baby cohort: n = 5,107). We used multiple imputation models with chained equations to generate 20 datasets for each cohort separately. The multiple imputation model included all confounding, exposure, and outcome variables used in the main and sensitivity analyses, along with several auxiliary variables: socio-economic position at study entry, state where resident, and parents’ migration history. We applied the population weight from the first wave to the imputation model, as this was available for all participants.^23^ Estimation models were then restricted to adolescents that were attending school at the time of the exposure (not imputed) (Kindergarten cohort: n = 3,917; Baby cohort: n = 3,359). We pooled the results from the imputed datasets together according to Rubin’s rules.

## Results

### Primary analyses

We analysed data from 3,200 adolescents (80·8% of participants that attended our baseline wave) in the Kindergarten cohort and 2,671 in the Baby cohort (79% of participants that attended our baseline wave) (Figure 1).

**Figure 1.**
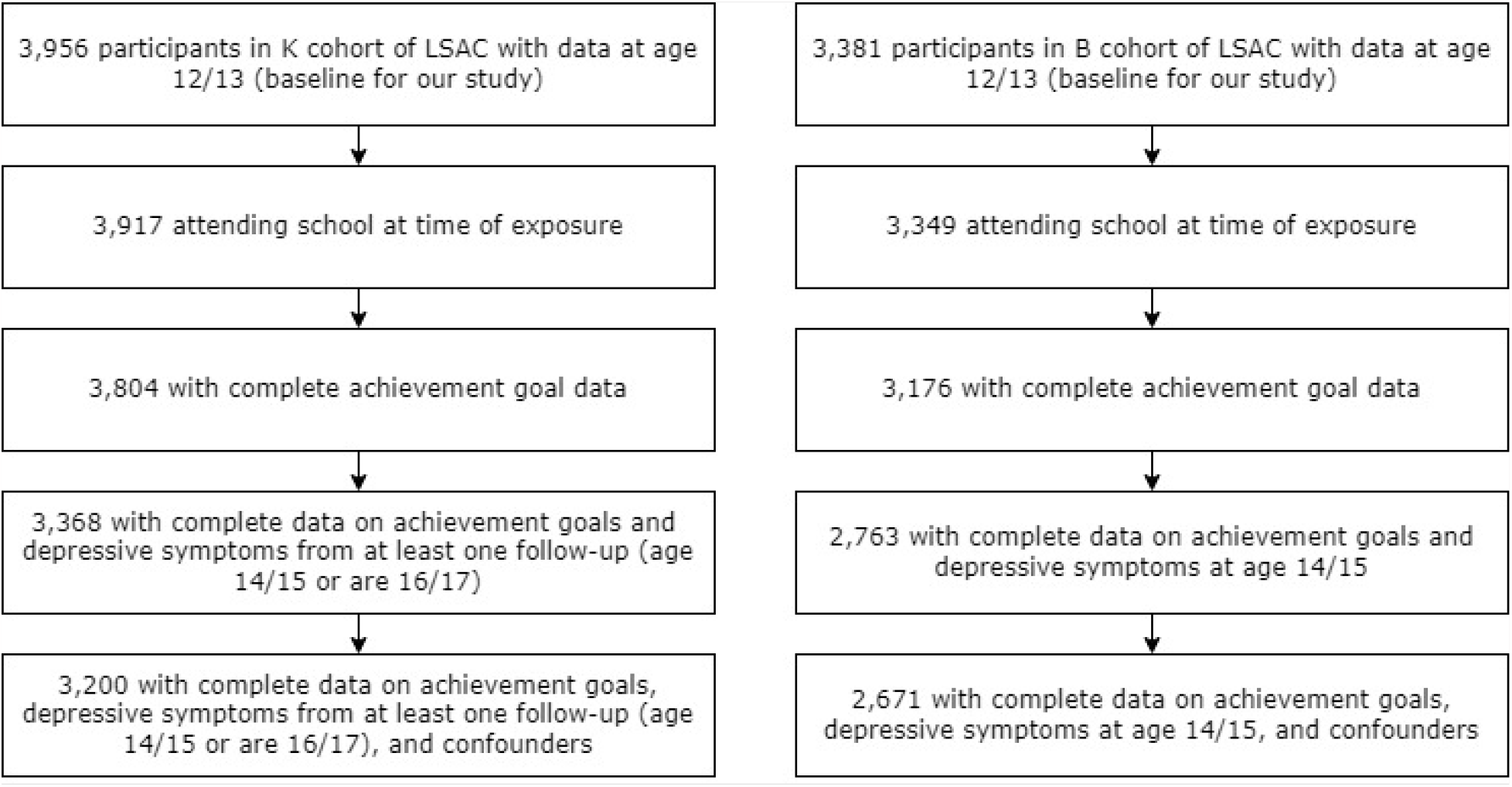
Study flowchart.

Characteristics of the samples according to AGQ subscale at age 12/13 scores are shown in Table 1. Of the AGQ subscales, mean scores were highest for mastery-approach and lowest for mastery-avoidance in each cohort.

**Table 1.** Characteristics of the sample with complete data, according to each AGQ subscales.

| Kindergarten cohort |  |  |  |  |
| --- | --- | --- | --- | --- |
|  | Mastery-approach | Mastery-avoidance | Performance-approach | Performance-avoidance |
|  | Mean (SD) | Mean (SD) | Mean (SD) | Mean (SD) |
| Total sample (n=3,200) | 5.56 (1.24) | 3.41 (1.59) | 4.64 (1.58) | 4.74 (1.50) |
| Sex |  |  |  |  |
| Female (n=1,585, 49.5%) | 5.61 (1.19) | 3.43 (1.60) | 4.43 (1.62) | 4.61 (1.49) |
| Male (n=1,615, 50.5%) | 5.52 (1.27) | 3.40 (1.57) | 4.84 (1.50) | 4.87 (1.50) |
| Two-parent household |  |  |  |  |
| No (n=448, 14%) | 5.39 (1.37) | 3.58 (1.62) | 4.54 (1.56) | 4.60 (1.47) |
| Yes (n=2,752, 86%) | 5.60 (1.21) | 3.38 (1.58) | 4.66 (1.58) | 4.77 (1.51) |
| SMFQ score at age 12/13: |  |  |  |  |
| Above cut-off (n=540, 16.9%) | 5.16 (1.36) | 4.19 (1.57) | 4.44 (1.59) | 4.69 (1.51) |
| Below cut-off (n=2,660, 83.1%) | 5.65 (1.19) | 3.25 (1.54) | 4.69 (1.57) | 4.75 (1.50) |
| Maternal K6 score at age 12/13 |  |  |  |  |
| Above cut-off (n=92, 2.9%) | 5.24 (1.56) | 3.85 (1.49) | 4.46 (1.60) | 4.86 (1.47) |
| Below cut-off (n=3,108, 97.1%) | 5.58 (1.23) | 3.40 (1.59) | 4.65 (1.58) | 4.74 (1.50) |
| Baby cohort |  |  |  |  |
|  | Mastery-approach | Mastery-avoidance | Performance-approach | Performance-avoidance |
|  | Mean (SD) | Mean (SD) | Mean (SD) | Mean (SD) |
| Total sample (n=2,671) | 5.51 (1.26) | 3.48 (1.58) | 4.66 (1.59) | 4.71 (1.53) |
| Sex |  |  |  |  |
| Female (n=1,310, 49%) | 5.58 (1.23) | 3.54 (1.59) | 4.48 (1.65) | 4.65 (1.56) |
| Male (n=1,361, 51%) | 5.44 (1.28) | 3.43 (1.57) | 4.83 (1.50) | 4.77 (1.50) |
| Two-parent household |  |  |  |  |
| No (n=371, 13.9%) | 5.26 (1.39) | 3.65 (1.56) | 4.54 (1.49) | 4.65 (1.44) |
| Yes (n=2,300, 86.1%) | 5.55 (1.23) | 3.46 (1.58) | 4.68 (1.60) | 4.72 (1.55) |
| SMFQ score at age 12/13: |  |  |  |  |
| Above cut-off (n=520, 19.5%) | 5.15 (1.41) | 4.06 (1.65) | 4.60 (1.62) | 4.83 (1.52) |
| Below cut-off (n=2,151, 80.5%) | 5.60 (1.21) | 3.34 (1.53) | 4.67 (1.58) | 4.69 (1.53) |
| Maternal K6 score at age 12/13 |  |  |  |  |
| Above cut-off (n=66, 2.5%) | 5.40 (1.38) | 3.82 (1.74) | 4.74 (1.55) | 4.98 (1.64) |
| Below cut-off (n=2,605, 97.5%) | 5.51 (1.26) | 3.47 (1.58) | 4.66 (1.59) | 4.71 (1.53) |

Participants with missing data in both cohorts had lower scores of mastery-approach goals, and higher scores of mastery-avoidance goals at age 12/13 compared to those in the complete cases samples (Supplementary Table 3). In the Kindergarten cohort, participants with missing data had higher levels of depressive symptoms at age 12/13. Participants with missing data were also more likely to live in a single parent household, have a lower score on the socio-economic composite variable, and have a mother with higher levels of depressive symptoms.

### Mastery goals

#### Kindergarten cohort

In univariable analyses, a 1-point increase in mastery-approach was associated with a −0·43 (95% CI −0·59 to −0·26) decrease in depressive symptoms (model one, table 2). Evidence of this association remained after all adjustments (−0·33 [95% −0·52 to −0·15]; model five, table 2). There was no evidence that the magnitude of the association varied by time-point (p value for interaction term: 0·206).

**Table 2.** Change in depressive symptoms at ages 14/15 and 16/17 (from linear multilevel model), per 1-point increase in achievement goals at age 12/13 (n=3,200); Kindergarten cohort.

| Model | Mastery goals |  |  |  | Performance goals |  |  |  |
| --- | --- | --- | --- | --- | --- | --- | --- | --- |
|  | Approach |  | Avoidance |  | Approach |  | Avoidance |  |
|  | Change in depression symptom severity score (95% CI) | p value | Change in depression symptom severity score (95% CI) | p value | Change in depression symptom severity (95% CI) | p value | Change in depression symptom severity (95% CI) | p value |
| Model one<sup>a</sup> | -0.43 (-.59 to -0.26) | <0.001 | 0.67 (0.54 to 0.80) | <0.001 | -0.16 (-0.29 to -0.02) | 0.021 | 0.15 (0.01 to 0.28) | 0.031 |
| Model two<sup>b</sup> | -0.49 (-0.68 to -0.3) | <0.001 | 0.67 (0.53 to 0.81) | <0.001 | -0.16 (-0.32 to 0.00) | 0.057 | 0.18 (0.02 to 0.34) | 0.030 |
| Model three<sup>c</sup>: | -0.49 (-0.68 to -0.3) | <0.001 | 0.67 (0.54 to 0.81) | <0.001 | -0.16 (-0.32 to 0.00) | 0.056 | 0.18 (0.01 to 0.34) | 0.033 |
| Model four<sup>d</sup> | -0.23 (-0.43 to -0.06) | 0.01 | 0.39 (0.25 to 0.53) | <0.001 | -0.16 (-0.32 to 0.00) | 0.045 | 0.21 (0.05 to 0.36) | 0.010 |
| Model five<sup>e</sup> | -0.33 (-0.52 to -0.15) | <0.001 | 0.35 (0.21 to 0.48) | <0.001 | -0.06 (-0.21 to 0.10) | 0.473 | 0.26 (0.11 to 0.41) | <0.001 |

<sup>a</sup>univariable. <sup>b</sup>model one plus all other AGQ subscales. <sup>c</sup>model two plus time variable. <sup>d</sup>model three adjusted for baseline depressive symptoms. <sup>e</sup>model four adjusted for all remaining confounders: mother's baseline depressive symptoms, gender, socio-economic position, number of parents in household. Interactions between AGQ subscales and time were added to model five; results for interaction terms are reported in the text.

A 1-point increase in mastery-avoidance was associated with a 0·67 (95% CI 0·54 to 0·80) increase in depressive symptoms in the univariable model (model one, table 2). Strong evidence of this association remained after adjustments (0·35 [95% 0·21 to 0·48]; model five, table 2). There was little evidence that this association varied by time-point (p value for interaction term: 0·070).

#### Baby cohort

In the univariable model, a 1-point increase in the mastery-approach subscale was associated with a −0·28 (95% CI −0·51 to −0·05) decrease in depressive symptoms at age 14/15 (model one, table 3). Strong evidence of this association remained after all adjustments (−0·29 [95% −0·54 to −0·03]; model five, table 3).

**Table 3.** Change in depressive symptoms at age 14/15, per 1-point increase in achievement goal orientations at age 12/13 (n=2,671); Baby cohort.

| Model | Mastery goals |  |  |  | Performance goals |  |  |  |
| --- | --- | --- | --- | --- | --- | --- | --- | --- |
|  | Approach |  | Avoidance |  | Approach |  | Avoidance |  |
|  | Change in depression symptom severity score (95% CI) | p value | Change in depression symptom severity score (95% CI) | p value | Change in depression symptom severity (95% CI) | p value | Change in depression symptom severity (95% CI) | p value |
| Model one<sup>a</sup> | -0.28 (-0.51 to -0.05) | 0.018 | 0.62 (0.44 to 0.81) | <0.001 | 0.04 (-0.15 to 0.22) | 0.688 | 0.11 (-0.08 to 0.30) | 0.269 |
| Model two<sup>b</sup> | -0.40 (-0.66 to -0.15) | 0.002 | 0.66 (0.47 to 0.86) | <0.001 | 0.09 (-0.14 to 0.31) | 0.444 | -0.05 (-0.28 to 0.19) | 0.701 |
| Model three<sup>c</sup>: | -0.18 (-0.44 to 0.07) | 0.163 | 0.49 (0.30 to 0.68) | <0.001 | 0.07 (-0.15 to 0.29) | 0.547 | -0.06 (-0.29 to 0.17) | 0.597 |
| Model four<sup>d</sup> | -0.29 (-0.54 to -0.03) | 0.027 | 0.44 (0.25 to 0.64) | <0.001 | 0.18 (-0.03 to 0.40) | 0.105 | -0.04 (-0.27 to 0.19) | 0.650 |

<sup>a</sup>univariable. <sup>b</sup>model one plus all other AGQ subscales. <sup>c</sup>model three adjusted for baseline depressive symptoms. <sup>d</sup>model four adjusted for all remaining confounders: mother's baseline depressive symptoms, gender, socio-economic position, number of parents in household.

A 1-point in increase in mastery-avoidance goals was associated with a 0·62 (95% CI 0·44 to 0·81) increase in depressive symptoms in the univariable model (model one, table 3). Strong evidence of this association remained after adjustments (0·44 [95% 0·25 to 0·64]; model five, table 3).

### Performance goals

#### Kindergarten cohort

In the univariable model, a 1-point increase in performance-approach goals was associated with a −0·16 (95% CI −0·29 to −0·02) decrease in depressive symptoms (model one, table 2). Evidence of this association was unaltered until the final model where it attenuated after adjusting for remaining confounders (−0·06 [95% −0·21 to 0·10]; model five, table 2). There was no evidence that the magnitude of the association varied by time-point (p=0·312).

A 1-point increase in performance-avoidance goals was associated with a 0·15 (95% CI 0·01 to 0·28) increase in depressive symptoms in the univariable model (model one, table 2). This association was unaltered until the final model where the effect estimate and the strength of the statistical evidence increased slightly (0·26 [95% 0·11 to 0·41]; model five, table 2). The magnitude of the association between performance-avoidance and depressive symptoms reduced as follow-up progressed (p value for interaction term: 0·026). When analysed in separate models, performance-avoidance goals were positively associated with depressive symptoms at both age 14/15 and age 16/17 in the fully adjusted models (table 4).

**Table 4.** Change in SMFQ depressive symptom severity score separately by time-point, per 1-point increase in performance-avoidance goals at age 12/13; Kindergarten cohort.

| Model | Outcome at 14/15 (n=3,068) |  | Outcome at 16/17 (n=2,686) |  |
| --- | --- | --- | --- | --- |
|  | Change in depression symptom severity (95% CI) | p value | Change in depression symptom severity (95% CI) | p value |
| Unadjusted | 0.28 (0.12 to 0.44) | <0.001 | 0.07 (-0.12 to 0.27) | 0.476 |
| Fully adjusted<sup>a</sup> | 0.38 (0.20 to 0.57) | <0.001 | 0.25 (0.02 to 0.48) | 0.034 |

<sup>a</sup>all other AGQ subscales, baseline depressive symptoms, mother's baseline depressive symptoms, gender, socio-economic position, number of parents in household.

#### Baby cohort

There was no evidence of an association between performance-approach goals and depressive symptoms at follow-up, in either the unadjusted (0·04 [95% CI −0·15 to −0·22]; model one, table 3) or fully adjusted models (0·18 [95% CI −0·03 to 0·40]; model four, table 3). Performance-avoidance goals had no association with future depressive symptoms at age 14/15 in either the unadjusted (0·11 [95% CI −0·08 to −0·30] model one, table 3) or fully adjusted models (−0·04 [95% CI −0·27 to 0·19] model four, table 3).

### Sensitivity analyses

The results of the sensitivity analyses are reported in the supplement. The association between mastery goals, and performance-approach goals with psychological distress measured with the K10 at age 18/19 were similar to the results from the main analyses (Supplementary Tables 4 & 5). However, there was no evidence for an association between performance-avoidance goals at age 12/13 (0·14 [95% CI: −0·13 to 0·40]) and 16/17 (−0·10 [95% CI: −0·40 to 0·19]) with psychological distress at age 18/19 respectively, whereas a positive association was found in the main analysis.

The addition of NAPLAN scores to the final model had minimal impact on the regression coefficients in the main analyses (Supplementary Table 6). In the multiple imputation analyses, results were similar to those from our complete case analysis (Supplementary Tables 7 to 9). However, in the multiple imputation analysis performance-approach goals at age 12/13 were positively associated with future depressive symptoms at age 14/15 in the Baby cohort in the fully adjusted model. For our primary analyses, we found a similar same pattern of association between the AGQ subscales and depression when using a binary depression variable in both the Baby and Kindergarten cohorts (Supplementary Table 10 to 12). However, a positive association between performance-avoidance goals at age 12/13 with depressive symptoms in the Kindergarten cohort was not found in the fully adjusted model using a binary depression outcome.

## Discussion

### Summary of findings

In two large nationally representative cohorts, we found consistent evidence that mastery goals were associated with subsequent depressive symptoms during adolescence. Higher mastery-approach goals were associated with lower levels of subsequent depressive symptoms, whilst higher mastery-avoidance goals was associated with higher subsequent depressive symptoms. We observed this pattern across a range of ages during secondary school.

Our findings in relation to performance goals were less consistent than for mastery goal. In the Kindergarten cohort, there was evidence of a positive association between performance-avoidance goals at age 12/13 and depressive symptoms at age 14/15 and 16/17, but no association with performance-approach. By the time the outcome was assessed at ages 18/19, there was no evidence of an association for any performance goals. These findings provided some support, albeit inconsistent, for hypotheses consistent with achievement goal theory, that higher performance goals are associated with higher depressive symptoms, at least during mid-(but not late) adolescence. However, in the Baby cohort, there was no evidence of an association with performance-approach or avoidance. These findings are in contrast to a previous longitudinal study which did not adjust for potential confounders, where higher levels of both performance-approach and performance-avoidance goals were associated with increased risk of depressive symptoms in Finish adolescents.^18^

### Strengths and limitations

We conducted the first large representative population-based cohort study to investigate associations between achievement goals and depressive symptoms. Use of two cohorts for replication is a strength of our study. Missing data is a limitation of most cohort studies and there was systematic attrition. However, we used multiple imputation to examine the impact of missing data and findings were predominantly consistent with our complete case analysis. Although we adjusted for a wide range of confounders, we are unable to rule out the possibility of residual confounding, for example due to personality or genetic factors.

### Mechanisms

There are several mechanisms that might explain why mastery-approach goals were associated with lower, and mastery-avoidance with higher, levels of subsequent depressive symptoms. Mastery goal orientation may be relevant to theories of psychological vulnerability to depression. Dysfunctional attitudes and negative attributions are modifiable belief systems found to increase the risk of depression in longitudinal population-based studies.^24,25^ Dysfunctional attitudes involve core beliefs such as ‘if I fail at my work then I am a failure as a person.’ Negative attributions for events are those that are internal (our fault), global (impacts all aspects of our world) and stable (will impact our future).^26^ By appraising failure as an opportunity to learn and grow, mastery-approach orientation might reduce dysfunctional attitudes and negative attributions and decrease vulnerability to depression.

Mastery-approach goals could also promote growth mindsets; core beliefs that abilities can be developed through practice.^12,27^ Growth mindsets could encourage adaptive ways of coping with stress and failure. There is evidence from cohort studies and RCTs that growth mindsets are associated with reduced levels of subsequent depressive symptoms.^28,29^ Conversely, mastery-avoidance goals could increase the risk of these experiences, which are potentially associated with increased risk of depressive symptoms.^26^

### Implications

Our findings support the hypothesis that mastery-approach goals are associated with lower levels of subsequent depressive symptoms in adolescents. If this relationship were causal, findings suggest that secondary school environments that emphasize learning, development, and personal growth (i.e., mastery goals) could reduce the risk of subsequent depressive symptoms during adolescence. School-based interventions to promote mastery goals could be evaluated in randomised controlled trials of preventative interventions for depressive symptoms.

## Data sharing

The data used in the study is available from the Australian Data Archive Dataverse: https://dataverse.ada.edu.au/dataverse.xhtml?alias=lsac

## Supporting information

The AGQ was completed at age 12/13 in each cohort, and again at age 16/17 in the Kindergarten cohort (Supplementary Table 1).

