## Supplementary material for "The association between academic achievement goals and adolescent depressive symptoms: a prospective cohort study in Australia": The AGQ was completed at age 12/13 in each cohort, and again at age 16/17 in the Kindergarten cohort (Supplementary Table 1).

| **Content** | **Description** | **Page number** |
| --- | --- | --- |
| Supplementary Table 1 | Details on the B and Kindergarten cohort and time-points when measures were collected | 2 |
| Supplementary Figure 1 | Directed Acyclic Graph showing the proposed causal relationship between achievement goal orientations and future depressive symptoms | 2 |
| Supplementary Table 2 | Correlation between achievement goal subscales | 3 |
| Supplementary Table 4 | Characteristics of the samples used for analyses compared with the samples excluded because of missing data. | 4 |
| Association between achievement goals and depressive symptoms at age 18/19 in the Kindergarten cohort |  | 5 |
| Supplementary Tables 5 & 6 | Sensitivity analysis with K10 outcome measure at age 18/19 | 6 |
| Supplementary Table 7 | Sensitivity analysis adjusting for NAPLAN scores | 7 |
| Supplementary Tables 8 to 10 | Sensitivity analyses using multiple imputation for missing data | 7, 8 |
| Supplementary Tables 11 to 13 | Sensitivity analysis of primary outcome using logistic regression | 9, 10 |
| STROBE checklist |  | 11 |

Supplementary Table 1. Details of each cohort, and when achievement goals (AGQ), depressive symptoms (sMFQ) and psychological distress (K10) were measured.

| Age | **Total sample size at each wave** | | **Measurement in each cohort** | | | | | |
| --- | --- | --- | --- | --- | --- | --- | --- | --- |
|  |  |  | **AGQ** | | **sMFQ** | | **K10** | |
|  | **K** | **B** | **K** | **B** | **K** | **B** | **K** | **B** |
| 12/13 | 3,956 | 3,381 | Yes | Yes | Yes | Yes | No | No |
| 14/15 | 3,537 | 3,127 | No | No | Yes | Yes | No | No |
| 16/17 | 3,089 | 2,017 | Yes | No | Yes | No | No | Yes (COVID-19) |
| 18/19 | 3,037 | - | Yes | - | No | - | Yes | - |

K: Kindergarten cohort; B: Baby cohort; AGQ: Achievement Goal Questionnaire; sMFQ: Short Mood and Feelings Questionnaire; K10: Kessler Psychological Distress Scale.

Yes: item was measured

No: Item was not measured

-: Cohort has not been followed-up at this age yet

Supplementary Figure 1. Directed Acyclic Graph showing the proposed causal relationship between achievement goal orientations and future depressive symptoms, including potential confounders available in the LSAC dataset. We have not included arrows between different confounders.

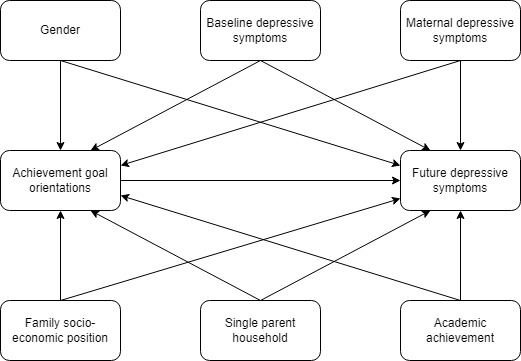

Supplementary Table 2. Correlation between achievement goal subscales.

| Kindergarten cohort – age 12/13 | | | |
| --- | --- | --- | --- |
|  | Mastery approach | Mastery avoidance | Performance approach |
| Mastery avoidance | 0·08 |  |  |
| Performance approach | 0·44 | 0·12 |  |
| Performance avoidance | 0·38 | 0·29 | 0·50 |
| Kindergarten cohort – age 16/17 | | | |
|  | Mastery approach | Mastery avoidance | Performance approach |
| Mastery avoidance | 0·22 |  |  |
| Performance approach | 0·51 | 0·14 |  |
| Performance avoidance | 0·39 | 0·34 | 0·54 |
| Baby cohort – age 12/13 | | | |
|  | Mastery approach | Mastery avoidance | Performance approach |
| Mastery avoidance | 0·12 |  |  |
| Performance approach | 0·42 | 0·17 |  |
| Performance avoidance | 0·38 | 0·34 | 0·54 |

Supplementary Table 3. Characteristics of the samples used for analyses compared with the samples excluded because of missing data.

|  | Kindergarten cohort | | | Baby cohort | | |
| --- | --- | --- | --- | --- | --- | --- |
| Characteristic | Analytical sample (n=3,200) | Excluded sample (n=717) | *p value* | Analytical sample (n=2,671) | Excluded sample (n=678) | *p value* |
| Sex |  |  |  |  |  |  |
| Female | 1,585 (49·5%) | 351 (46·4%) |  | 1,310 (49%) | 328 (48·4%) |  |
| Male | 1,615 (50·5%) | 405 (53·6%) | 0·125 | 1,361 (51%) | 350 (51·6%) | 0·454 |
| Two-parent household |  |  |  |  |  |  |
| No | 448 (14%) | 226 (30·1%) |  | 371 (13·9%) | 146 (23·4%) |  |
| Yes | 2,752 (86%) | 525 (69·9%) | <0·001 | 2,300 (86·1%) | 478 (76·6%) | <0·001 |
| Mastery approach at age 12/13 | 5·56 (1·24) | 5·35 (1·40) | <0·001 | 5·51 (1·26) | 5·34 (1·35) | 0·0046 |
| Mastery avoidance at age 12/13 | 3·41 (1·59) | 3·64 (1·64) | <0·001 | 3·48 (1·58) | 3·68 (1·54) | 0·0093 |
| Performance approach at age 12/13 | 4·64 (1·58) | 4·60 (1·59) | 0·557 | 4·66 (1·59) | 4·67 (1·49) | 0·877 |
| Performance avoidance at age 12/13 | 4·74 (1·50) | 4·69 (1·51) | 0·471 | 4·71 (1·53) | 4·67 (1·49) | 0·577 |
| SMFQ at age 12/13 | 3·97 (5·21) | 4·52 (5·80) | 0·0167 | 4·30 (6·05) | 4·76 (6·28) | 0·127 |
| Socio-economic position at age 12/13 | 0·082 (0·982) | -0·371 (0·997) | <0·001 | 0·058 (0·994) | -0·249 (0·996) | <0·001 |
| Maternal K6 score at age 12/13 | 9·06 (3·63) | 9·61 (4·37) | 0·0018 | 9·01 (3·47) | 9·67 (4·12) | <0·001 |

Data are mean (SD) or n (%). Analytical samples are those with complete data on exposure, confounders, and depressive symptoms at a least one time-point. Both the analytical and excluded sample were attending school at age 12/13.

**Association between achievement goals and psychological distress at age 18/19 in the Kindergarten cohort**

We used a sample of 2,406 adolescents in the Kindergarten cohort (60·8% of participants that attended our baseline wave) with complete data for secondary analyses of AGQ scores at age 12/13 and psychological distress at age 18/19 measured with the K10. For the AGQ at age 16/17 and psychological distress at age 18/19 measured with the K10, we used a sample of 1,919 adolescents (56·8% of participants that attended our baseline wave) with complete data.

The association between mastery goals and psychological distress at age 18/19 were similar to the association with depressive symptoms at age 14/15 and 16/17 found in the main analyses. In the fully adjusted models, a 1-point increase in mastery approach goals at age 12/13 and 16/17 was associated with a -0·49 (95% CI -0·81 to -0·16) and a -0.68 decrease (95% -1·02 to -0·35) in psychological distress at age 18/19 respectively (Supplementary Table 4). A 1-point increase in mastery avoidance goals at age 12/13 and 16/17 was associated with a 0·53 (95% CI 0·30 to 0·76) and a 0.85 increase (95% 0·61 to 1·10) in psychological distress at age 18/19 respectively after adjustment for confounders (Supplementary Table 4).

No association was found between performance approach goals at age 12/13 (0.07 [95% CI: -0·19 to 0·34]) and 16/17 (0·10 [95% CI: -0·16 to 0·35]) with psychological distress at age 18/19 respectively (Supplementary Table 5), replicating the findings in the main analyses. However, performance avoidance goals at age 12/13 (0·14 [95% CI: -0·13 to 0.40]) and 16/17 (-0·10 [95% CI: -0·40 to 0·19]) were not associated with psychological distress at age 18/19 respectively (Supplementary Table 5). In the main analysis a positive association was found with depressive symptoms.

Supplementary Table 4. Change in K10 psychological distress severity score at age 18/19, per 1-point increase in mastery approach and mastery avoidance goals at age 12/13 (n=2,406); Kindergarten cohort.

|  | Mastery goals | | | | Performance goals | | | |
| --- | --- | --- | --- | --- | --- | --- | --- | --- |
|  | Approach | | Avoidance | | Approach | | Avoidance | |
|  | Change in depression symptom severity score (95% CI) | p value | Change in depression symptom severity score (95% CI) | p value | Change in depression symptom severity (95% CI) | p value | Change in depression symptom severity (95% CI) | p value |
| Model one: univariable association | -0·52 (-0·81 to -0·23) | <0·001 | 0·83 (0·61 to 1·06) | <0·001 | -0·13 (-0·35 to 0·10) | 0·280 | 0·10 (-0·13 to 0·33) | 0·376 |
| Model two: model one plus all other AGQ subscales | -0·59 (-0·91 to 0·26) | <0·001 | 0·85 (0·62 to 1·09) | <0·001 | -0·08 (-0·35 to 0·20) | 0·587 | 0·07 (-0·20 to 0·35) | 0·599 |
| Model three: model two adjusted for baseline depressive symptoms | -0·38 (-0·7 to -0·05) | 0·022 | 0·58 (0·35 to 0·82) | <0·001 | -0·7 (-0·34 to 0·20) | 0·605 | 0·08 (-0·18 to 0·35) | 0·532 |
| Model four: model three adjusted for all remaining confounders* | -0·49 (-0·81 to -0·16) | 0·003 | 0·53 (0·30 to 0·76) | <0·001 | 0·07 (-0·19 to 0·34) | 0·575 | 0·14 (-0·13 to 0·40) | 0·304 |

* mother’s baseline depressive symptoms, gender, socio-economic position, number of parents in household.

Supplementary Table 5. Change in K10 psychological distress severity score at age 18/19, per 1-point increase in mastery approach and mastery avoidance goals at age 16/17 (n=1,919); Kindergarten cohort.

|  | Mastery goals | | | | Performance goals | | | |
| --- | --- | --- | --- | --- | --- | --- | --- | --- |
|  | Approach | | Avoidance | | Approach | | Avoidance | |
|  | Change in depression symptom severity score (95% CI) | p value | Change in depression symptom severity score (95% CI) | p value | Change in depression symptom severity (95% CI) | p value | Change in depression symptom severity (95% CI) | p value |
| Model one: univariable association | -0·56 (-0·89 to -0·24) | <0·001 | 1·17 (0·93 to 1·42) | <0·001 | -0·22 (-0·47 to 0·02) | 0·075 | 0·10 (-0·19 to 0·40) | 0·494 |
| Model two: model one plus all other AGQ subscales | -0·92 (-1·27 to -0·58) | <0·001 | 1·37 (1·12 to 1·62) | <0·001 | -0·02 (-0·30 to 0·25) | 0·881 | -0·05 (-0·37 to 0·26) | 0·738 |
| Model three: model two adjusted for baseline depressive symptoms | -0·61 (-0·93 to -0·28) | <0·001 | 0·95 (0·70 to 1·20) | <0·001 | 0·03 (-0·22 to 0·28) | 0·806 | -0·11 (-0·41 to 0·18) | 0·456 |
| Model four: model three adjusted for all remaining confounders* | -0·68 (-1·02 to -0·35) | <0·001 | 0·85 (0·61 to 1·10) | <0·001 | 0·10 (-0·16 to 0·35) | 0·459 | -0·10 (-0·40 to 0·19) | 0·492 |

* mother’s baseline depressive symptoms, gender, socio-economic position, number of parents in household.

Supplementary Table 6. Results of the sensitivity analyses with NAPLAN scores added to the final model.

|  | Mastery goals | | | | Performance goals | | | |
| --- | --- | --- | --- | --- | --- | --- | --- | --- |
|  | Approach | | Avoidance | | Approach | | Avoidance | |
|  | Change in depression symptom severity score (95% CI) | p value | Change in depression symptom severity score (95% CI) | p value | Change in depression symptom severity score (95% CI) | p value | Change in depression symptom severity score (95% CI) | p value |
| Kindergarten cohort |  |  |  |  |  |  |  |  |
| Final model adjusting for Year 7 NAPLAN scores*(n=2,979) | -0·36 (-0·55 to -0·17) | <0·001 | 0·36 (0·22 to 0·51) | <0·001 | -0·08 (-0·24 to 0·08) | 0·339 | 0·29 (0·14 to 0·45) | <0·001 |
| Baby cohort |  |  |  |  |  |  |  |  |
| Final model adjusting for Year 7 NAPLAN scores* (n=2,384) | -0·27 (-0·56 to 0·00) | 0·056 | 0·45 (0·24 to 0·67) | <0·001 | 0·15 (-0·08 to 0·39) | 0·204 | -0·06 (-0·31 to 0·19) | 0·650 |

*adjusted for all other AGQ subscales, baseline depressive symptoms, mother’s baseline depressive symptoms, gender, socio-economic position, number of parents in household

Supplementary Table 7. Change in depressive symptoms at ages 14/15 and 16/17 (from linear multilevel model), per 1-point increase in achievement goals at age 12/13 (imputed sample; n=3,917); Kindergarten cohort.

|  | Mastery goals | | | | Performance goals | | | |
| --- | --- | --- | --- | --- | --- | --- | --- | --- |
|  | Approach | | Avoidance | | Approach | | Avoidance | |
|  | Change in depression symptom severity score (95% CI) | p value | Change in depression symptom severity score (95% CI) | p value | Change in depression symptom severity (95% CI) | p value | Change in depression symptom severity (95% CI) | p value |
| Model one: univariable association | -0·43 (-0·59 to -0·26) | <0·001 | 0·61 (0·48 to 0·74) | <0·001 | -0·17 (-0·30 to -0·04) | 0·012 | 0·12 (-0·01 to 0·25) | 0·075 |
| Model two: model one plus all other AGQ subscales | -0·51 (-0·70 to -0·32) | <0·001 | 0·62 (0·49 to 0·76) | <0·001 | -0·15 (-0·31 to 0·00) | 0·053 | 0·18 (0·02 to 0·34) | 0·024 |
| Model three: model two plus time variable | -0·51 (-0·70 to -0·32) | <0·001 | 0·62 (0·49 to 0·76) | <0·001 | -0·15 (-0·31 to 0·00) | 0·053 | 0·18 (0·02 to 0·34) | 0·034 |
| Model four: model three adjusted for baseline depressive symptoms | -0·25 (-0·44 to -0·6) | 0·01 | 0·35 (0·21 to 0·49) | <0·001 | -0·15 (-0·3 to 0·00) | 0·046 | 0·21 (0·06 to 0·36) | 0·007 |
| Model five: model four adjusted for all remaining confounders* | -0·33 (-0·52 to -0·15) | <0·001 | 0·31 (0·17 to 0·45) | <0·001 | -0·05 (-0·19 to 0·1) | 0·537 | 0·26 (0·11 to 0·41) | 0·023 |

*gender, socio-economic position, number of parents in household

Supplementary Table 8. Change in SMFQ depressive symptom severity score separately by time-point, per 1-point increase in performance avoidance goals at age 12/13; Kindergarten cohort imputed sample.

|  | Outcome at 14/15 (n=3,917) | | Outcome at 16/17 (n=3,917) | |
| --- | --- | --- | --- | --- |
|  | Change in depression symptom severity (95% CI) | p value | Change in depression symptom severity (95% CI) | p value |
| Unadjusted | 0·75 (0·60 to 0·90) | <0·001 | 0·04 (-0·26 to 0·24) | 0·41 |
| Fully adjusted* | 0·34 (0·15 to 0·52) | <0·001 | 0·22 (-0·02 to 0·46) | 0·07 |

*all other AGQ subscales, baseline depressive symptoms, mother’s baseline depressive symptoms, gender, socio-economic position, number of parents in household

Supplementary table 9. Change in depressive symptoms at age 14/15, per 1-point increase in achievement goals at age 12/13 (imputed sample; n=3,349); Baby cohort.

|  | Mastery goals | | | | Performance goals | | | |
| --- | --- | --- | --- | --- | --- | --- | --- | --- |
|  | Approach | | Avoidance | | Approach | | Avoidance | |
|  | Change in depression symptom severity score (95% CI) | p value | Change in depression symptom severity score (95% CI) | p value | Change in depression symptom severity score (95% CI) | p value | Change in depression symptom severity score (95% CI) | p value |
| Model one: univariable association | -0·30 (-0·53 to -0·07) | 0·010 | 0·58 (0·39 to 0·76) | <0·001 | 0·05 (-0·14 to 0·24) | 0·576 | 0·06 (-0·15 to 0·26) | 0·581 |
| Model two: model one plus all other AGQ subscales | -0·44 (-0·69 to -0·19) | <0·001 | 0·63 (0·44 to 0·83) | <0·001 | 0·15 (-0·08 to 0·37) | 0·21 | -0·10 (-0·35 to 0·15) | 0·443 |
| Model three: model two adjusted for baseline depressive symptoms | -0·21 (-0·46 to 0·05) | 0·110 | 0·45 (0·26 to 0·65) | <0·001 | 0·13 (-0·09 to 0·35) | 0·029 | -0·10 (-0·35 to 0·14) | 0·408 |
| Model four: model three adjusted for all remaining confounders* | -0·32 (-0·57 to -0·07) | 0·012 | 0·40 (0·20 to 0·60) | <0·001 | 0·24 (0·02 to 0·46) | 0·037 | -0·07 (-0·31 to 0·18) | 0·599 |

*gender, socio-economic position, number of parents in household

Supplementary Table 10. Odds ratios for depression per 1-point increase in achievement goals at age 12/13; (n=3,200) Kindergarten cohort.

|  | Mastery goals | | | | Performance goals | | | |
| --- | --- | --- | --- | --- | --- | --- | --- | --- |
|  | Approach | | Avoidance | | Approach | | Avoidance | |
|  | Odds ratio (95% CI) | p value | Odds ratio (95% CI) | p value | Odds ratio (95% CI) | p value | Odds ratio (95% CI) | p value |
| Model one: univariable association | 0·84 (0·78 to 0·91) | <0·001 | 1·29 (1·21 to 1·38) | <0·001 | 0·94 (0·88 to 1·01) | 0·070 | 1·03 (0·96 to 1·10) | 0·380 |
| Model two: model one plus all other AGQ subscales | 0·83 (0·76 to 0·91) | <0·001 | 1·29 (1·21 to 1·38) | <0·001 | 0·96 (0·89 to 1·04) | 0·304 | 1·03 (0·95 to 1·11) | 0·484 |
| Model three: model two plus time variable | 0·83 (0·76 to 0·91) | <0·001 | 1·29 (1·21 to 1·38) | <0·001 | 0·96 (0·89 to 1·04) | 0·304 | 1·03 (0·95 to 1·11) | 0·491 |
| Model four: model three adjusted for baseline depressive symptoms | 0·88 (0·80 to 0·96) | 0·005 | 1·22 (1·14 to 1·31) | <0·001 | 0·96 (0·89 to 1·04) | 0·304 | 1·05 (0·96 to 1·12) | 0·368 |
| Model five: model four adjusted for all remaining confounders* | 0·84 (0·77 to 0·93) | <0·001 | 1·21 (1·13 to 1·29) | <0·001 | 1 (0·92 to 1·08) | 0·991 | 1·06 (0·98 to 1·15) | 0·121 |

* mother’s baseline depressive symptoms, gender, socio-economic position, number of parents in household.

Supplementary Table 11. Odds ratios for depression separately by time-point, per 1-point increase in performance avoidance goals at age 12/13; Kindergarten cohort.

|  | Outcome at 14/15 (n=3,068) | | Outcome at 16/17 (n=2,686) | |
| --- | --- | --- | --- | --- |
|  | Odds ratio (95% CI) | p value | Odds ratio (95% CI) | p value |
| Unadjusted | 1·08 (1·03 to 1·15) | 0·004 | 1·00 (0·96 to 1·06) | 0·759 |
| Fully adjusted* | 1·17 (1·09 to 1·27) | <0·001 | 1·05 (0·98 to 1·12) | 0·169 |

*all other AGQ subscales, baseline depressive symptoms, mother’s baseline depressive symptoms, gender, socio-economic position, number of parents in household

Supplementary Table 12. Odds ratios for depression at age 14/15 per 1-point increase in achievement goals at age 12/13; (n=2,671) Baby cohort.

|  | Mastery goals | | | | Performance goals | | | |
| --- | --- | --- | --- | --- | --- | --- | --- | --- |
|  | Approach | | Avoidance | | Approach | | Avoidance | |
|  | Odds ratio (95% CI) | p value | Odds ratio (95% CI) | p value | Odds ratio (95% CI) | p value | Odds ratio (95% CI) | p value |
| Model one: univariable association | 0·89 (0·83 to 0·95) | <0·001 | 1·21 (1·15 to 1·29) | <0·001 | 0·97 (0·92 to 1·03) | 0·331 | 1·00 (0·95 to 1·06) | 0·849 |
| Model two: model one plus all other AGQ subscales | 0·86 (0·79 to 0·93) | <0·001 | 1·25 (1·18 to 1·26) | <0·001 | 0·99 (0·93 to 1·07) | 0·921 | 0·98 (0·91 to 1·05) | 0·511 |
| Model three: model two adjusted for baseline depressive symptoms | 0·92 (0·84 to 0·99) | 0·039 | 1·18 (1·12 to 1·26) | <0·001 | 0·99 (0·93 to 1·07) | 0·839 | 0·96 (0·89 to 1·04) | 0·303 |
| Model four: model three adjusted for all remaining confounders* | 0·88 (0·81 to 0·96) | 0·003 | 1·18 (1·10 to 1·26) | <0·001 | 1·03 (0·96 to 1·11) | 0·407 | 0·97 (0·90 to 1·05) | 0·397 |

* mother’s baseline depressive symptoms, gender, socio-economic position, number of parents in household.

STROBE Statement.

|  | | Item No | Recommendation | Page |
| --- | --- | --- | --- | --- |
| **Title and abstract** | | 1 | (*a*) Indicate the study’s design with a commonly used term in the title or the abstract | Title |
|  |  |  | (*b*) Provide in the abstract an informative and balanced summary of what was done and what was found | Abstract |
| Introduction | | | | |
| Background/rationale | | 2 | Explain the scientific background and rationale for the investigation being reported | Introduction |
| Objectives | | 3 | State specific objectives, including any prespecified hypotheses | Introduction |
| Methods | | | | |
| Study design | | 4 | Present key elements of study design early in the paper | Study design and participants |
| Setting | | 5 | Describe the setting, locations, and relevant dates, including periods of recruitment, exposure, follow-up, and data collection | Study design and participants |
| Participants | | 6 | (*a*) *Cohort study*—Give the eligibility criteria, and the sources and methods of selection of participants. Describe methods of follow-up  *Case-control study*—Give the eligibility criteria, and the sources and methods of case ascertainment and control selection. Give the rationale for the choice of cases and controls  *Cross-sectional study*—Give the eligibility criteria, and the sources and methods of selection of participants | Study design and participants |
|  |  |  | (*b*) *Cohort study*—For matched studies, give matching criteria and number of exposed and unexposed  *Case-control study*—For matched studies, give matching criteria and the number of controls per case | N/A |
| Variables | | 7 | Clearly define all outcomes, exposures, predictors, potential confounders, and effect modifiers. Give diagnostic criteria, if applicable | Outcome/Exposure/ Potential confounders |
| Data sources/ measurement | | 8* | For each variable of interest, give sources of data and details of methods of assessment (measurement). Describe comparability of assessment methods if there is more than one group | Outcome/Exposure/ Potential confounders |
| Bias | | 9 | Describe any efforts to address potential sources of bias | Sensitivity analyses |
| Study size | | 10 | Explain how the study size was arrived at | Study design and participants |
| Quantitative variables | | 11 | Explain how quantitative variables were handled in the analyses. If applicable, describe which groupings were chosen and why |  |
| Statistical methods | | 12 | (*a*) Describe all statistical methods, including those used to control for confounding | Statistical analysis |
|  |  |  | (*b*) Describe any methods used to examine subgroups and interactions | N/A |
|  |  |  | (*c*) Explain how missing data were addressed | Sensitivity analyses |
|  |  |  | (*d*) *Cohort study*—If applicable, explain how loss to follow-up was addressed  *Case-control study*—If applicable, explain how matching of cases and controls was addressed  *Cross-sectional study*—If applicable, describe analytical methods taking account of sampling strategy | N/A |
|  |  |  | (*e*) Describe any sensitivity analyses | Sensitivity analyses |
| Results | | | | |
| Participants | 13* | (a) Report numbers of individuals at each stage of study—eg numbers potentially eligible, examined for eligibility, confirmed eligible, included in the study, completing follow-up, and analysed | | Figure 1 |
|  |  | (b) Give reasons for non-participation at each stage | | Figure 1 |
|  |  | (c) Consider use of a flow diagram | | Figure 1 |
| Descriptive data | 14* | (a) Give characteristics of study participants (eg demographic, clinical, social) and information on exposures and potential confounders | | Table 1 |
|  |  | (b) Indicate number of participants with missing data for each variable of interest | | Figure 1 |
|  |  | (c) *Cohort study*—Summarise follow-up time (eg, average and total amount) | | N/A |
| Outcome data | 15* | *Cohort study*—Report numbers of outcome events or summary measures over time | | N/A |
|  |  | *Case-control study—*Report numbers in each exposure category, or summary measures of exposure | | N/A |
|  |  | *Cross-sectional study—*Report numbers of outcome events or summary measures | | N/A |
| Main results | 16 | (*a*) Give unadjusted estimates and, if applicable, confounder-adjusted estimates and their precision (eg, 95% confidence interval). Make clear which confounders were adjusted for and why they were included | | Results – Primary analyses |
|  |  | (*b*) Report category boundaries when continuous variables were categorized | | N/A |
|  |  | (*c*) If relevant, consider translating estimates of relative risk into absolute risk for a meaningful time period | | N/A |
| Other analyses | 17 | Report other analyses done—eg analyses of subgroups and interactions, and sensitivity analyses | | Results – Sensitivity analyses |
| Discussion | | | | |
| Key results | 18 | Summarise key results with reference to study objectives | | Discussion - Summary of findings |
| Limitations | 19 | Discuss limitations of the study, taking into account sources of potential bias or imprecision. Discuss both direction and magnitude of any potential bias | | Discussion – Strengths and limitations |
| Interpretation | 20 | Give a cautious overall interpretation of results considering objectives, limitations, multiplicity of analyses, results from similar studies, and other relevant evidence | | Discussion – Implications |
| Generalisability | 21 | Discuss the generalisability (external validity) of the study results | | Discussion – Strengths and limitations |
| Other information | | | | |
| Funding | 22 | Give the source of funding and the role of the funders for the present study and, if applicable, for the original study on which the present article is based | | Abstract |
